# Hormone Receptor Expression and Disease Prognosis in High-Grade Serous Ovarian Cancer

**DOI:** 10.1101/2023.04.21.23288934

**Authors:** Leah V. Dodds, Alex P Sanchez-Covarrubias, Ramlogan Sowamber, Anca Milea, Andre Pinto, Yuguang Ban, Matthew Schlumbrecht, Patricia A Shaw, Sophia HL George

**Affiliations:** Department of Obstetrics, Gynecology and Reproductive Sciences, Division of Gynecologic Oncology, University of Miami Miller School of Medicine, Miami, FL, USA; Medical Scientist Training Program, Prevention Sciences Graduate Program, University of Miami Miller School of Medicine, Miami, FL, USA; Sylvester Comprehensive Cancer Center; Cancer Biology Graduate Program, University of Miami Miller School of Medicine, Miami, FL, USA; Princess Margaret Cancer Center, University Health Network, Ontario, Canada; Department of Medicine, Internal Medicine Residency Program, University of Miami Miller School of Medicine, Miami, FL, USA; Department of Pathology, University of Miami Miller School of Medicine, Miami, FL, USA; Department of Epidemiology and Population Health Sciences, Division of Biostatistics and Bioinformatics, University of Miami Miller School of Medicine, Miami, FL, USA

## Abstract

ER and PR regulate growth and differentiation in normal ovaries and fallopian tubes and in HGSC transformation and progression. Higher PR expression was associated with improved survival outcomes, while high ER expression was associated with worse survival in patients with HGSC. Here, we show that patients with ER+PR+ tumors have longer overall survival and confirm the role of PR as a prognostic marker of survival and response to chemotherapy. Gene expression analysis demonstrated up-regulation of the ATM signaling pathway in the ER+PR+ subgroup when compared to ER+PR− tumors. Up-regulation of interferon alpha, beta and gamma signaling, and antigen presentation pathways were identified in ER+PR− compared to ER−PR+. In summary, this study elucidated that the genomic and transcriptomic signatures related to ER/PR status in HGSC have clinical prognostic value.

## Introduction

Epithelial ovarian cancer (EOC) is a heterogeneous group of diseases with multiple histological subtypes and is broadly divided into a dualistic model based on morphological, molecular genetics, and clinical features^1–3^. There are at least five subtypes of EOC: low grade serous carcinoma (LGSC), high-grade serous carcinoma (HGSC) (often referred to as fallopian tube cancers), endometrioid, mucinous and clear cell carcinomas^4^. HGSC is the most common and aggressive histotype, accounting for approximately 75% of ovarian carcinomas and 85% of the cancer-related deaths and it is characterized by significant genomic instability, immune cell infiltration and inflammation^4, 5^. HGSC commonly express the estrogen receptor and most cases originate from the fallopian tube^6^. Cyclic exposure to estrogen, found in the follicular fluid increases inflammation of the fallopian tube through reactive oxidative species (ROS) and subsequently induces DNA damage stress^7^. Conditions that increase lifetime ovulation events, including infertility, low parity, delayed menopause, and early menarche, are epidemiologically linked to increased HGSC risk^8, 9^. Therefore, hormonal milieu and the presence of the estrogen and progesterone receptors in the fallopian tube epithelium (FTE) suggest and support a role for these hormones in disease pathogenesis.^9–13^

The presence of both estrogen receptor alpha (ER) and progesterone receptor (PR) expression in epithelial ovarian tumors, like l LGSC tumors, has been shown to predict favorable response to estrogen antagonism^14^. PR is a classic target of ER and a biomarker of ER function^15^. LGSC tumors have a higher rate of ER/PR expression than HGSC^16, 17^. In *recurrent* LGSC, antiestrogen therapies (aromatase inhibitors and selective estrogen receptor modulator) have low overall response rates (9-11%) but can yield stable disease (>70%) and improve progression free survival^14, 18^. In contrast, only 10-26% of platinum resistant recurrent HGSC patients will experience stable disease when treated with hormonal/antiestrogen therapy^18–21^.

The biological basis for a role of estrogen in ovarian carcinogenesis includes stimulation of growth signaling pathways including ERK/MAPK, p38/MAPK, PI3K/AKT, and PLC/PKC through growth factors and cytokines^22–24^. A now well-established stepwise ovarian cancer initiation model whereby fallopian tube epithelia acquires genomic alterations that then promote the transition to serous tubal intraepithelial carcinoma (STIC)^25–28^, prompted us to investigate the relationship between ER/PR in HGSC development, including the role of ER expression in the presence and absence of PR. Further, we investigated the relationship of ER/PR expression on progression free survival and overall survival and explored the impact of ER/PR driven transcriptomics and genomics in HGSC.

## Materials and Methods

### Case collection

The study protocols for collection of biological specimens and clinical information for all patients was approved by the University Health Network (UHN) Research Ethics Board (#02-0882) and University of Miami (UM) Institutional Review Board (#2015-1022). Snap frozen (n=75) HGSC tissue samples from a previously published paper^29^ and formalin-fixed paraffin embedded tissues (FFPE) (n=523) were retrospectively selected from the UHN and UM biobanks. A total of 375 of the 523 cases had ER/PR immunohistochemical data and was used for further analysis - 148 cases were excluded because of missing clinical or immunohistochemical data. Clinical data was available for 193 of the 375 cases and included age, date of diagnosis, stage, surgical debulking, first line chemotherapy, date of recurrence, date of death, date of last follow-up and vital status. Cases that were snap frozen (n=75) were selected for copy number alteration analysis.

### Immunohistochemistry

Tissue microarrays (TMAs) were constructed using tissue from patients with HGSC and patients with no known mutation, otherwise considered normal, on a semiautomated TMArrayer (Pathology Devices, Inc., San Diego, CA, USA), as described in previous publications^29^. In short, 8 TMAs with 0.6mm tumor punches and duplicate or triplicate cores were used. Standard immunohistochemistry was performed on each TMA with antibodies listed in **Supplement S1**. Stained slides were scanned using the ScanScope XT slide scanner (Aperio Technologies, Inc., Leica, Buffalo Grove, Il, USA) to create digital images at 40x magnification. The percentage of cells positive for expression was determined by quantifying ER and PR staining using a nuclear algorithm as previously described (Spectrum Plus, Image Analysis Toolbox, and TMALab II (Aperio Technologies Inc, Vista, CA) ^9, 30, 31^. The optimal cutoff for ER and PR expression is unclear^32^. For this study, ER and PR nuclei positivity was described as ≥ 5% nuclei positivity based on commonly used thresholds of 1%-9%^33, 34^. IHC was also performed with ≥ 10% nuclei positivity cutoff according to some clinical guidelines^34, 35^. Intensity levels were not considered for this study. Two FTE TMAs were used as previously described^29^ and a cohort of 15 STIC cases with adnexal normal FTE and HGSC was also reviewed for ER and PR expression by IHC^11, 29, 36^. Protein expression for duplicated and triplicated TMA cores were averaged before subsequent analysis. Image analysis of Ki67, CD3, CD8, CD68 and FOXP3 was performed as previously reported^11^. All images were annotated to include epithelium and exclude stroma.

### Statistical Analysis

Wilcoxon rank-sum was used for continuous variables in nonparametric distributions. Chi-square testing (or Fisher’s Exact, when appropriate) was used to analyze associations between categorical variables. Progression-free survival (PFS) was defined as the time elapsed between diagnosis and recurrence. Overall survival (OS) was defined as the time elapsed between diagnosis and death by disease or date of last follow-up. Survival analysis was performed using the Kaplan-Meier method on OS and PFS. Associations within groups broken down by ER and/or PR status were assessed by the Mantel-Haenszel log-rank test. Univariable and multivariable Cox proportional hazards regression analyses were performed to assess the effect of explanatory variables on OS. To assess the influence PR status as an independent predictor of OS, two clinical scenarios were modeled in the multivariable cox regression analysis. Results were reported as hazard ratios (HR) with 95% confidence intervals (95%CI). Stepwise backwards multivariable regression analyses included covariates with p < 0.05 from the univariable models (model 1) or strong clinically relevant covariates (model 2). All tests were two-sided, with significance set at p < 0.05. Statistical analysis was performed using STATA IC 17 (StataCorp, College Station, TX) and R 4.1.2 (Foundation for Statistical Computing, Vienna, Austria). The R packages survival^37^ and survminer^38^ were used to calculate and plot survival data.

### Transcriptomic Analysis

Our previously published cohort of 75 HGSC tumors (GSE10971 and GSE28044)^29, 36^ was used. Data analysis was performed using PARTEK v6.0 software (St. Louis, MO, USA). Data for each probe was preprocessed and normalized using the Robust Means Algorithm (RMA), followed by a median centered normalization for each gene across all samples. Analysis was performed on log base 2 transformed data. All gene lists generated were determined based on a fold change of 2+ and statistical significance of (FDR) q<0.05. Supervised hierarchical clustering was performed to determine the molecular phenotypes associated with clinically distinct groups based on ER/PR protein expression. Ingenuity Pathway Analysis (IPA) was used to visualize the interactive networks of genes found to be significantly represented and to determine the biological processes and molecular functions of these genes. Gene Set Enrichment Analysis (GSEA) (http://www.broadinstitute.org/gsea/index.jsp) was performed using MSigDB (C2:CP, ver.5.1) database for each gene set collection or using gene sets curated based on published data. Gene set size filters (min=15, max=5000) included 980 / 1330 gene sets in the analysis. All GSEA analyses in this study used statistically significant (p<0.05) enrichment scores derived from whole transcriptome data.

### Copy Number Alteration (CNA) analysis

A well characterized previously published cohort^29^ of 75 HGSC samples were genotyped using Affymetrix Genome-Wide Human SNP Array 6.0 (Santa Clara, CA, USA), according to the manufacturer’s instructions. From these cases, we compared 70 cases at gene regions and 64 cases at segment regions. Single Nucleotide Polymorphism (SNP) data was analyzed by importing CEL files into PARTEK Genomics Suite 7.0 (St. Louis, MO, USA)^29, 36^. Using multi-array average, quintile normalization and quality assessment via Principal Component Analysis (PCA), the data were corrected for background errors. CNA analysis for unpaired samples was performed using the reference file distributed by PARTEK to detect total copy number (CN) gains/losses. Specifically, genes and segment regions with CNAs were detected using the Genomic Segmentation algorithm available in PARTEK. Cutoffs were chosen for the following calls: >2.3 for CN gains, and >1.7 for CN losses. Using the R package ComplexHeatmap^39^, we plotted selected oncogenes, and tumor suppressor genes involved in the homologous recombination pathway based on the TCGA report of genes associated with epithelial ovarian cancer. In addition, in the segment analysis, the top 20 differentially CN gains/losses across groups were plotted similarly using the same package.

## Results

### Hormonal receptor expression confers differential survival outcomes

A total of 375 FFPE tissues were selected for ER/PR expression analyses. Using TMAs of fallopian tube epithelium, fimbriated sections and tumor tissue, we determined the proportion of HGSC tumors with expression of ER and PR. HGSC tumors were predominantly ER+ (76%, 285/375) and PR− (61%, 228/375) (**Fig. 1A, B**). Additionally, four subgroups, based on ER and PR expression, were identified: ER+PR− (41.9%, 157/375), ER+/PR+ (34.1%, 128/375), ER−PR− (18.9%, 71/375) and ER−PR+ (5.1% 19/375), (**Fig. 1C**). A subset of 197 cases had complete clinical data, including survival data. A summary of the clinicopathologic characteristics of these cases by ER/PR expression is found in **Table 1**. ER−PR+ patients had the youngest age at diagnosis 52.3 (+/− 10.1) compared to other groups (p=0.026). Fifty-three percent (53%) of the patients were optimally debulked and 90% received first line platinum/taxane chemotherapy.

**Figure 1:**
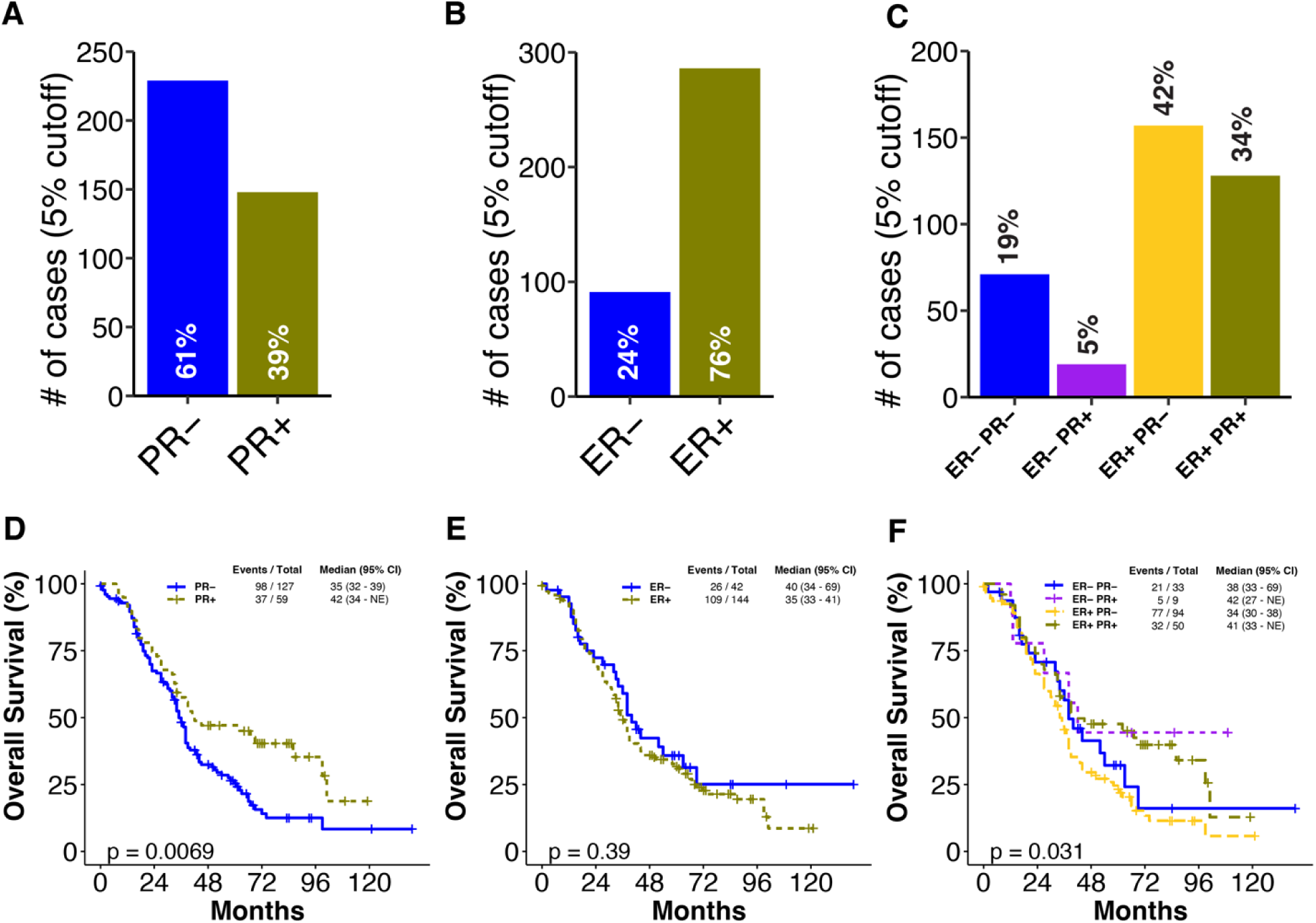
Top: Clinical characteristics of ER/PR subgroups. **A.** Quantification of 375 representative sections of HGSC stained with PR. PR expression is determined using a cut-off of 5%. Cases with greater than 5% positive cell staining are PR+. Total number of cases positive for PR is represented as a percentage. **B**. Representation of the number of cases with ER expression**. C.** Four subgroups (ER−PR− (n=71), ER−PR+(n=19), ER+PR− (n=157) and ER+PR+(n=128)) are analyzed for both ER and PR expression. A representation of the number of cases belonging to each subgroup using a 5% expression cut-off. Bottom: Overall Survival analysis by ER/PR protein expression. **D.** PR+ cases are associated with increased overall survival compared to samples that are PR− (p=0.0069). **E.** Overall survival analysis of ER+ vs ER− cases. ER+ cases do not show significant difference in survival outcome compared to ER− cases (p=0.39) **F**. Overall survival months for ER/PR subgroups. ER−PR+ cases confer increased overall survival compared to other subgroups (p=0.031), pairwise comparison showed differences in OS between ER+PR+ and ER+PR− groups only (p=0.008).

**Table 1.** Cohort Characteristics by ER/PR sub-groups at 5% expression cutoff **(n=197)**

| Variable | ER-/PR-<br>(n=34) | ER-/PR+<br>(n=9) | ER+/PR-<br>(n=100) | ER+/PR+<br>(n=54) | p<br>value |
| --- | --- | --- | --- | --- | --- |
| **Age in years (n=192)** |  |  |  |  |  |
| Mean (SD) | 60.4 (12.8) | 52.3 (10.1) | 67.8 (9.8) | 57.3 (13.3) | 0.026 |
| **Debulking status (n=182)** |  |  |  |  | 0.142 |
| Optimal | 16 (50) | 5 (62.5) | 44 (46.8) | 32 (66.7) |  |
| Suboptimal | 16 (50) | 3 (37.5) | 50 (53.2) | 16 (33.3) |  |
| **Family History of Breast or Ovarian Cancer (n= 150)** |  |  |  |  | 0.531 |
| Yes | 13 (50) | 7 (77.8) | 47 (60.3) | 23 (62.2) |  |
| No | 13 (50) | 2 (22.2) | 31 (39.7) | 14 (37.8) |  |
| **Chemotherapy (n=177)** |  |  |  |  | 0.660 |
| Platinum only | 5 (15.6) | 0 (0) | 8 (9.2) | 5 (10.2) |  |
| Platinum + Taxane | 27 (84.4) | 9 (100) | 79 (90.8) | 44 (89.8) |  |
| **Number of cycles (n=165)** |  |  |  |  | 0.648 |
| <6 | 3 (10.7) | 0 | 7 (8.4) | 2 (4.4) |  |
| ≥6 | 25 (89.3) | 9 (100) | 76 (91.6) | 43 (95.6) |  |
| **Stage (n=193)** |  |  |  |  | 0.013 |
| I – II | 5 (15.6) | 1 (11.1) | 4 (4) | 10 (19.2) |  |
| III - IV | 28 (84.4) | 8 (88.9) | 95 (96) | 42 (80.8) |  |

Survival analyses showed that women with PR+ tumors had better OS (median OS 42 months (95% confidence interval (CI), 34 - not estimable (NE))) compared to women with PR− tumors (median OS 35 months (95% CI, 32–38)) (p=0.0069, **Fig. 1D**). At 10% cutoff PR status maintained statistical significance **(Sup. S2E).** However, this trend was not observed in ER expression at any cutoff (**Fig. 1E, Sup. S2E**). OS across the combined categories resulted in a better OS in ER−PR+ tumors (median OS 42 months (95% CI, 27 – NE)) while the ER+PR− had the worse OS (median OS 34 (96% CI, 30-38)) (p=0.031, **Fig. 1F**). PFS analysis at 5% cutoff did not show statistically significant differences for ER+/− (p=0.47), PR+/− (p=0.54) nor the combined ER/PR subgroups (p=0.8) (**Sup. S2A-C**). Other non-significant OS and PFS analyses are included in **Sup. S2.**

A Cox regression univariable analysis showed an association of PR+ tumors with longer OS compared to PR− tumors (HR 1.69; 95%CI, 1.15 - 2.48) (p=0.008, Table S3); furthermore, pairwise comparison across 4 different ER/PR subgroups showed shorter OS in ER+ PR− when compared to ER+ PR+ (HR 1.76; 95%CI, 1.16 – 2.66) with no other pairwise comparison being significant. Other variables associated with higher mortality included suboptimal debulking status (HR 2.21; 95%CI, 1.54 – 3.17); not reporting a family history of Breast and/or Ovarian cancer (HR 1.55; 95%CI, 1.03-2.33); receiving a first course chemotherapy with platinum only (HR 1.75; 95%CI 1.01 – 3.07); receiving less than 6 cycles of chemotherapy (HR 5.24; 95%CI 2,78 – 9.85) and being diagnosed at an advance stage (HR 3.14; 95%CI 1.52 – 6.46). Model 1 included statistically significant variables in the univariable analysis and model 2 included clinically relevant variables known to strongly influence OS: debulking status, type of chemotherapy and number of cycles received, in addition to PR status. Because the majority of HGSC cases are diagnosed at a late stage, stage was not included in model 2. In multivariable analysis, for model 1, receipt of chemotherapy with platinum only (p=0.006) and receipt of less than 6 cycles (p=0.046) were independently associated with OS, whereas for model 2, debulking status was added as an independently associated predictor (p=0.004). Progesterone receptor status was not associated with OS in any of the multivariable models (**Sup. S3**).

### Differential gene expression analysis of HGSC tumors indicate differences amongst four ER/PR subgroups

In normal FTE and STIC cases, we sought to determine how early changes in ER/PR expression occur (**Fig. 2A**). In this analysis, FTE samples included the fallopian tube with some cases having fimbria where ER and PR expression were found in both ciliated and non-ciliated cell types^40^. Each STIC case was stained for TP53, Ki67, ER and PR. In premenopausal women without HGSC, the proportion of ER+ and PR+ cells in FTE varied by ovarian cycle status with more ER+ cells in the follicular phase (pre-ovulation/proliferative phase, 47.5%) compared to the luteal phase (post-ovulation/secretory phase, 38.3%; p=0.01) (**Sup. S4**).The proportion of ER+ cells between FTE and HGSC (38.5% vs. 22.8%, respectively, p=0.04) indicated a difference in expression levels, however in comparing ER expression levels in STIC (n=13) to both FTE (n= 9) and HGSC (n=13), a larger difference was noted indicating an increased level of ER expression in the lesion. (**Fig. 2B**). Moreover, PR+ expression levels decreased from FTE (17.4%) to STIC (16.0%) to HGSC (7.2%) (p= 0.005) (**Fig. 2B**).

**Figure 2:**
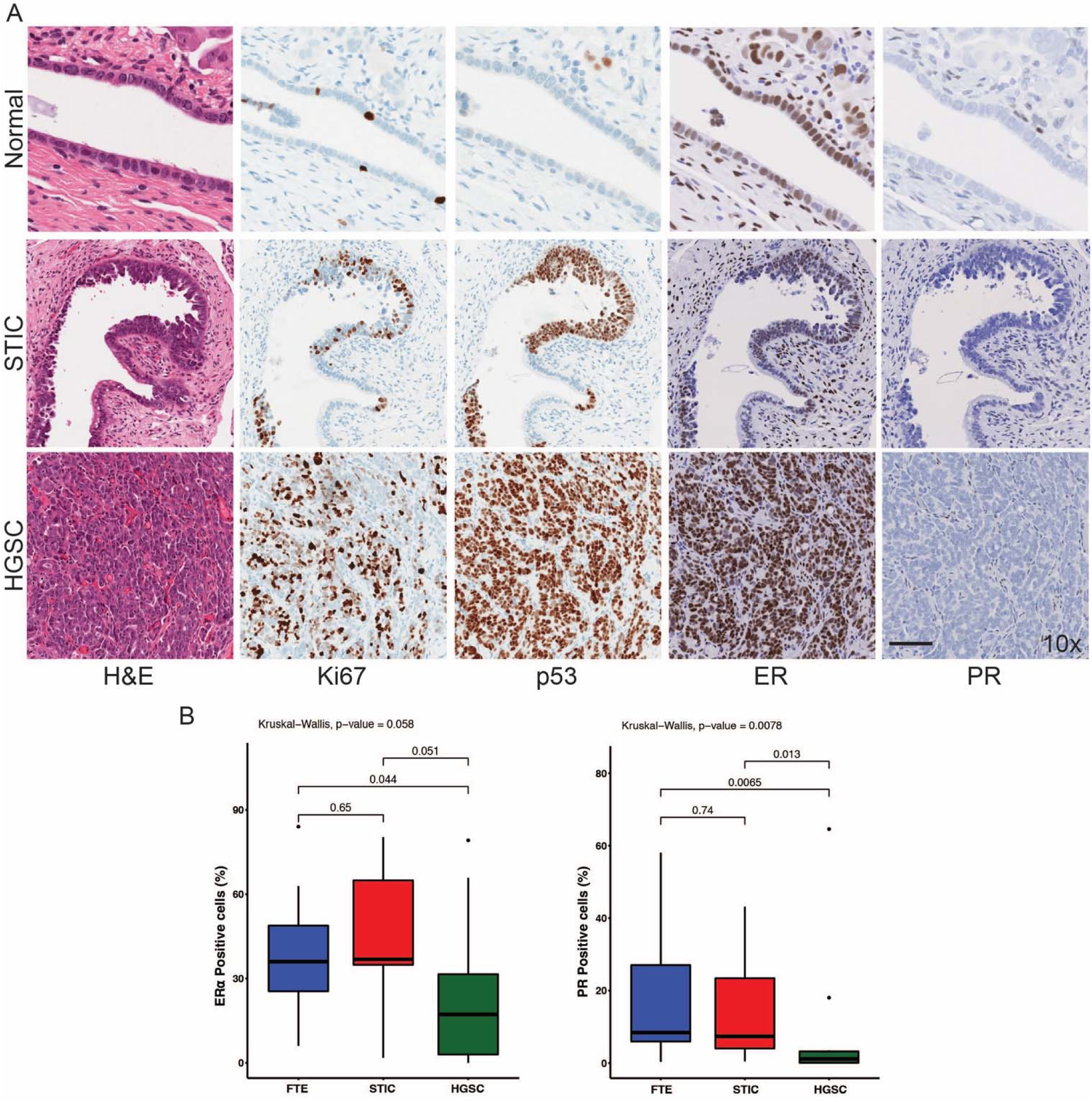
PR expression decreases in HGSC compared to normal fallopian tube tissue. **A**. Visual representation of ER, PR, Ki67 and p53 protein expression by immunohistochemistry. Sections of normal FTE tissue, serous tubal intraepithelial lesions (STIC) and HGSC tissue sections. Tissue morphology is determined by p53 and Ki67 (marker of proliferation). Positive protein expression, highlighted by darker stained cells is quantified using a custom nuclear algorithm (x10). **B.** Quantification of ER and PR protein expression in normal FTE (n=13), STIC (n=9) and HGSC (n=13). In ER positive cells, there was significant expression between FTE and HGSC in (p=0.04) and no other significant differences were found between STICS and other morphologies. In PR positive cells, significant differences were found between FTE and HGSC (p=0.007) and between STIC and HGSC (p=0.01).

To evaluate the effect of ER and PR on gene expression levels of FTE, we used a subset of 45 randomly selected HGSC tumors from the 375 cases and performed gene expression analysis for tumors classified into four subgroups, based on a 5% cutoff for ER and PR: ER−PR− (n=8), ER−PR+ (n=3), ER+PR− (n=18), ER+PR+ (n=13). 354 genes were differentially expressed (DEGs) between the four subgroups (p<0.005) (**Sup. S5**). Unsupervised hierarchical clustering showed distinct gene expression profiles of ER−PR+ compared to the three other subgroups (**Sup. S6**). Pairwise comparison showed that 152 genes were differentially expressed between ER+PR− and ER−PR+; 130 genes were differentially expressed between ER−PR+ and ER−PR− and 6 genes were differentially expressed between ER+PR− and ER+PR+.

The top DEGs between ER+PR− vs ER−PR+ included: SMOC1 (downregulated in ER+PR−, FC=-10.2, p=1.50 x 10^−22^), SLC4A4 (downregulated in ER+PR−, FC=-6.62, p=-2.06 x 10^−11^) and KLK8 (upregulated in ER+PR−, FC=21.9, p=9.37 x 10^−11^). The top DEGs between ER−PR+ and ER−PR− included also SMOC1 (upregulated in ER−PR+, FC=9.31, p=1.33 x 10^−18^), SLC4A4 (upregulated in ER−PR+, FC=7.99, p=2.56 x 10^−10^) and ZNF117 (upregulated in ER−PR+, FC=5.84, p=2.51 x 10^−9^). The top DEGs between ER+PR+ and ER+PR− included: SNUPN (downregulated in ER+PR+, FC=-2.13, p=2.33 x 10^−5^), OR2A20P/OR2A9P (downregulated in ER+PR+, FC=-2.42, p=1.59 x 10^−4^) and ASB9 (upregulated in ER+PR+, FC=3.25, p=1.04 x 10^−3^). These comparisons suggest a role for PR in driving specific transcriptional programming in HGSC. Other pairwise comparisons are reported in **Sup. S5**.

### Pathway analysis of PR driven transcriptional programs show differences with varying ER expression

We performed pathway analysis on differentially expressed genes between ER/PR subgroups to identify PR-driven gene transcriptional pathways and cellular programs in HGSC. A comparison between ER+PR− and ER−PR+ groups showed an upregulation of 85% of genes belonging to the antigen presenting pathway in the ER+PR− subgroup (p=0.00002) (**Sup. S7A, B**). Top differentially regulated pathways included phagosome maturation (TUBB3, LAMP2, NCF2), crosstalk between dendritic cells and natural killer cells (CD40, FAS, IL18) and dendritic cell maturation (CREB1, JAK2, STAT1) (**Sup. S7A-B, Sup. S8**). A comparison of ER− PR− and ER−PR+ showed the pathways regulated solely by PR, which included adaptive immune pathways such as allograft rejection signaling (TNF, CD40, B2M) (p=1.14 x 10^−5^) and the innate immune system which included the crosstalk between dendritic cells and natural killer cells (CD40, CD86, FSCN3) (p=3.30×10^−5^) (**Sup. S7D-E, Sup. S9**). Additionally, a comparison of ER+PR− and ER+PR+ subgroups, from which the OS survival difference was significant, showed PR driven gene transcriptional programs in the presence of ER, which included the ATM signaling pathway (ATF2, ATM, H2AX) (z-score=0.277, p = 7.18 x 10^−3^) as the most differentially regulated pathway, with >60% of the genes upregulated in the ER+PR+ subgroup (**Fig. 3A-B, Sup. S10**).

**Figure 3.**
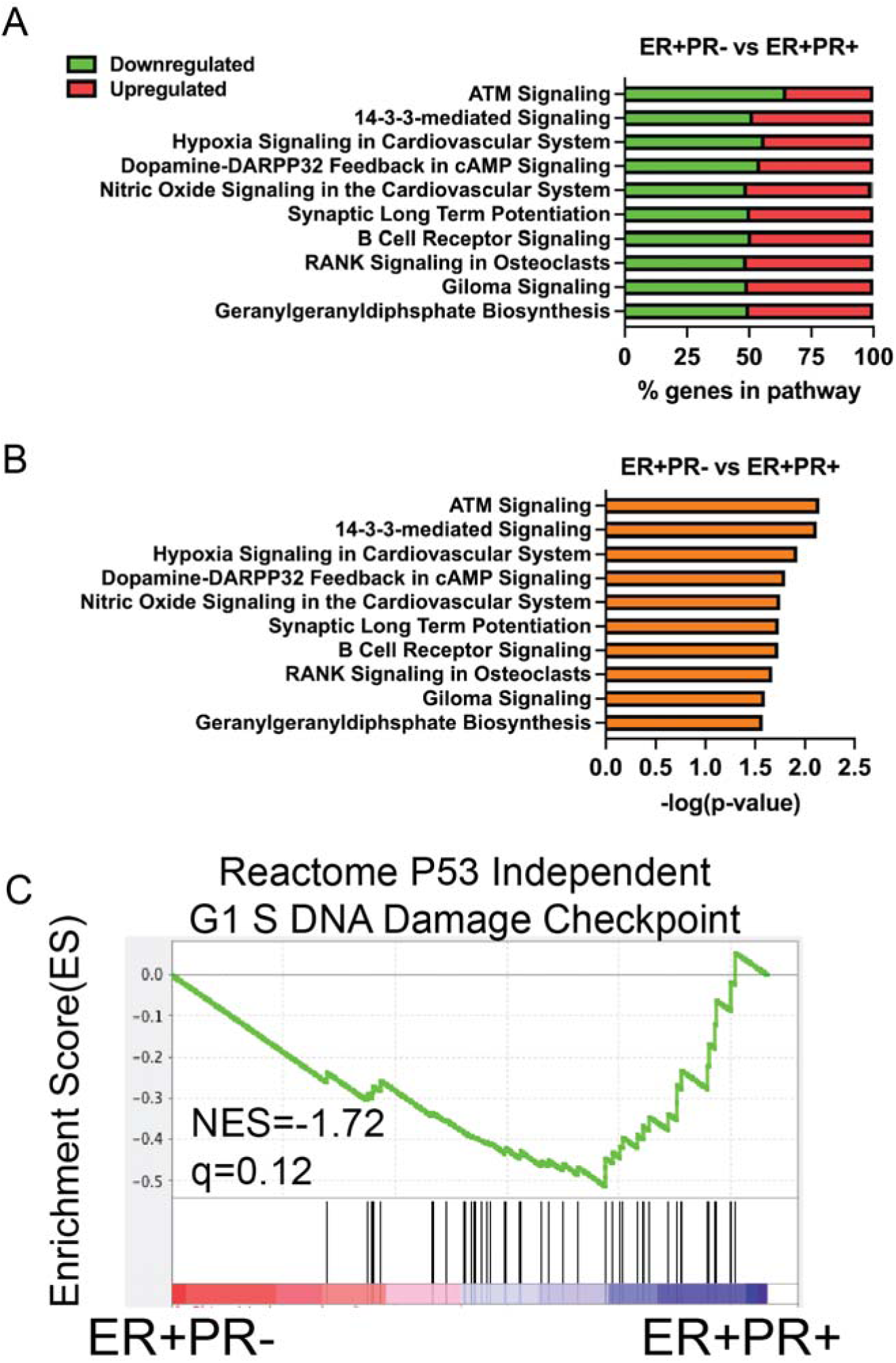
Comparison of ER signaling in the presence and absence of PR. **A.** A comparison between ER+PR− vs ER+PR+ shows percentage of genes upregulated and downregulated in the presence of ER. In the absence of PR, ER downregulates more than 50% of genes involves in the ATM signaling pathway. **B.** Top pathways identified in a comparison between ER+PR− and ER+PR+ **C**. Enrichment plot for a comparison of ER+PR− and ER+PR+ shows DNA damage checkpoint genes enriched (NES = −1.72, q =0.12).

Gene set enrichment analyses (GSEA) performed on two subgroup comparisons, indicative of PR signaling (ER−PR− vs ER−PR+; ER+PR− vs ER+PR+), showed enriched gene sets belonging to the DNA repair pathway and the p53 dependent G1/S DNA damage response when comparing ER+PR− vs ER+PR+ (**Fig. S7F, 3C**). Core enriched genes included BRCA1, BRIP1, MDC1 and LIG4, RAD51 and RAD52. GSEA performed on ER−PR+ vs ER+PR− showed an upregulation of adaptive and innate immune pathways in ER+PR− tumors. Specific genes upregulated in ER+PR− tumors included interferon alpha/beta (PTPN1, RNASEL, IFNA6; FDR<10^−5^, NES=-2.55), interferon gamma signaling (NCAM1, CAMK2D, PTPN1; FDR<10^−5^, NES=-2.46), cytokine signaling (NCAM1, SOCS2, PLCG1;FDR<10^−5^, NES=-2.33), antigen processing and presentation (HSPA1 and 6 family, NFYZ, CREB1; FDR=3.77 x 10^−5^, NES=-2.15), and IL12 pathways (PP3CA, PP3R1, MAPK14; FDR=2.71 x 10^−5^, NES=-2.17) (**Fig. S7C**).

Due to an enrichment of immune related pathways and gene signatures observed in ER−PR+ vs ER+PR−, we evaluated tumor infiltrating lymphocytes (TILs) and macrophages within the ER/PR subgroups (**Fig. 4A**). TMAs were stained for CD3, CD8, CD68 and FOXP3 immune markers and assessed using automated image analysis (**Fig. 4B**). There were no significant differences in CD3+, CD8+ and FOXP3+ cells across the different categories of ER/PR status (**Fig. 4C**). A significantly higher proportion of CD68+ immune cells were found in the ER+ group relative to the ER− group (p=0.03) (**Fig. 4D**).

**Figure 4.**
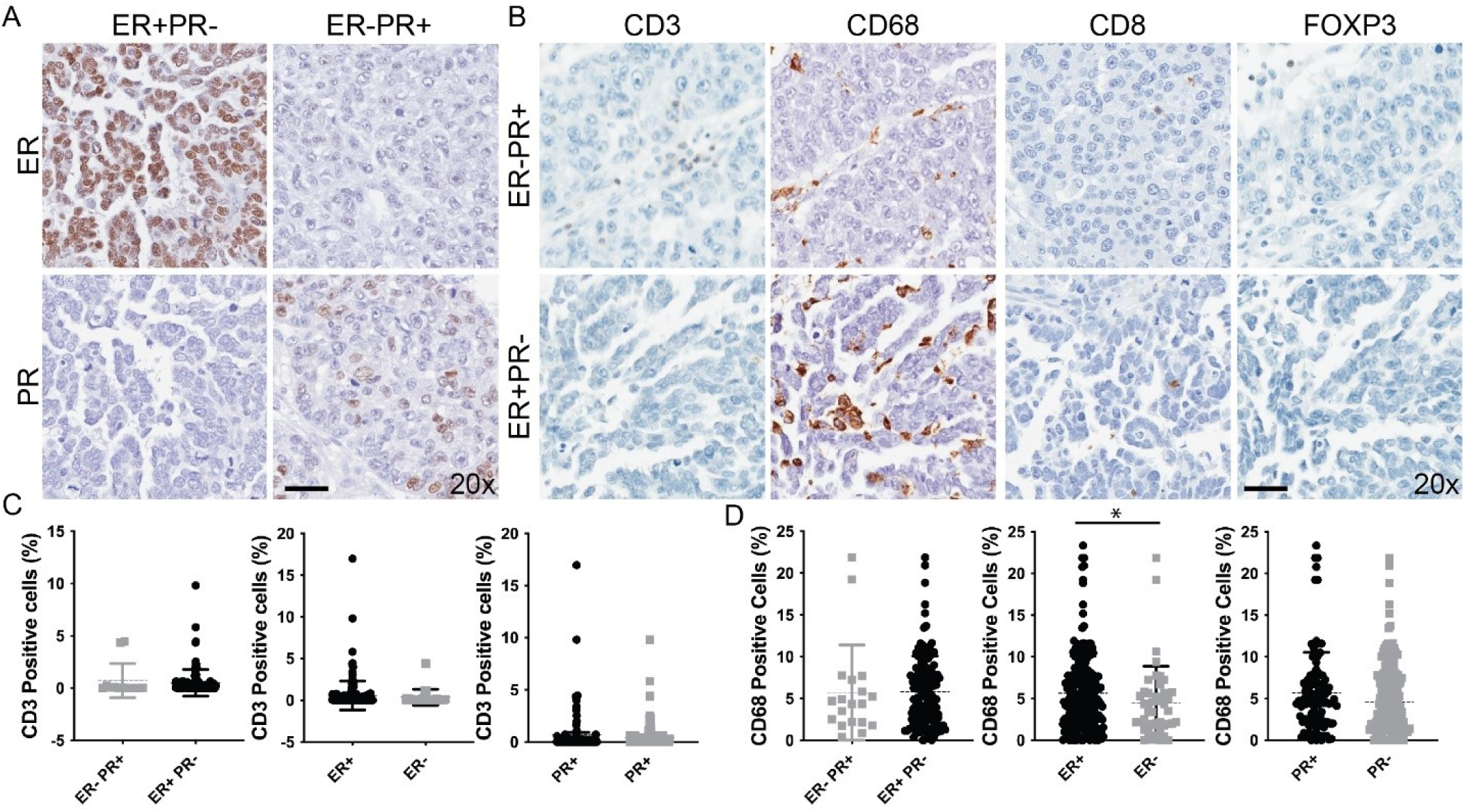
CD68 is enriched in PR+ cases. **A.** Sections of HGSC stained for ER and PR across ER+PR− and ER−PR+ subgroups. Positive cells are identified by brown nuclear staining (x20). B. CD3, CD68, CD8, FOXP3 staining of ER+PR− and ER−PR+ cases. Positive cells are identified by brown nuclear staining (x20). **C.** Quantification of CD3 positive cells using a custom nuclear algorithm for identification of positive CD3 cells**. D.** Quantification of CD68 positive cells shows more positivity in the ER+PR− subgroup. Higher percentage of CD68+ cells are found in the ER+ group compared to ER− group (p<0.05).

### Copy Number Alterations (CNA) Analysis and chromosomal segment analysis of ER/PR subgroups shows loss of MYC and genomic alteration of chromosomal region 22q11.22 in ER−PR+

The data of 75 snap frozen HGSC from our cohort was used to generate an Oncoplot showing CNAs in MYC, PIK3CA, CCNE1, PALB2, BRCA1, and BRCA2 genes which have been shown to be amplified or deleted in HGSC and linked to disease pathogenesis^41–44^(**Fig. 5A**). The MYC gene was lost (1 copy or less) in 86% (6/7) of cases in the ER−PR+ group compared to 59% (17/29) in the ER+PR− group (p=0.38). The tumor suppressor PALB2 was amplified in 34% (10/29) of cases in the ER+PR− group compared to 0% of cases in the ER− PR+ (p=0.15). BRCA1 showed a similar pattern with amplification in 57% (4/7) of cases in the ER−PR+ vs amplification of 34% (10/29) of cases in the ER+PR− (p=0.39). ESR1 was amplified in 8.3% (4/48) of ER+ cases compared to 9% (2/22) of ER− cases (p=1); PGR was deleted in 4.6% (2/44) of PR− cases compared to 0% of cases in the PR+ (p=0.34). It is unlikely that high ER protein expression in HGSC is due to amplification of the ESR1 gene, and low PR expression due to deletion of the PGR gene. Top copy number variations of oncogenes specific to HGSC that were amplified included MECOM (86%, 60/70) and MYC (74%, 52/70) across the entire cohort (**Sup. S11A**). Top copy number deletions were found in tumor suppressor genes TP53 (70%, 49/70), BRCA1 (60%, 42/70) and WWOX (54%,38/70) across all samples. (**Sup. S11B**).

**Figure 5.**
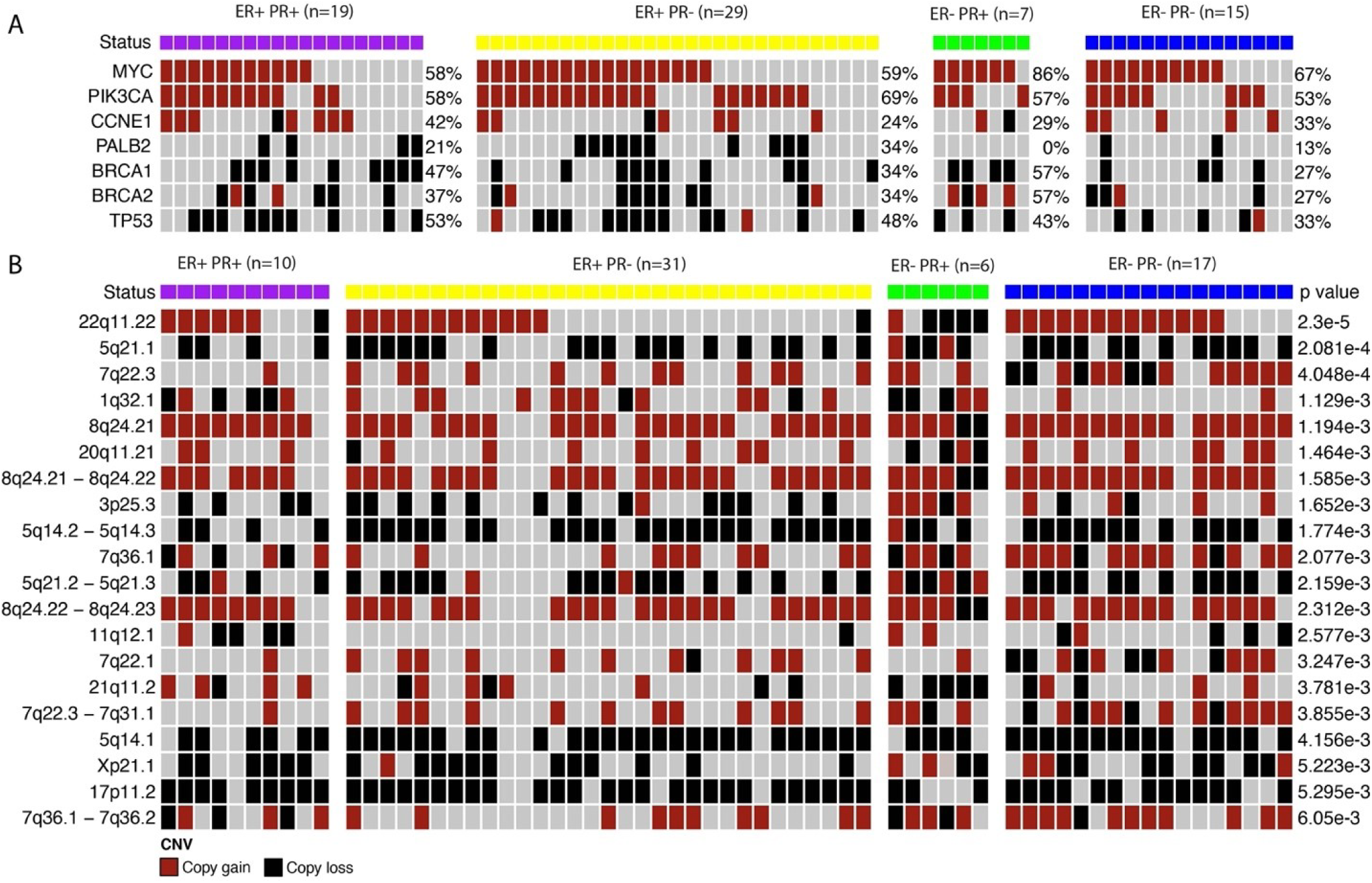
A. Copy number variation gene analysis. Selected genes involved in the homologous recombination pathway were selected across four groups (n=70). Selected oncogenes associated with ovarian cancer based on the TCGA (MYC, PICK3CA and CCNE1) presented copy gains uniformly in the cohort. Similarly, selected tumor suppressor genes involved in the ovarian cancer based on TCGA (PALB2, BRCA1, BRCA2, TP53). **B.** Copy number variation segment analysis. Top 20 segments differential CNV gains and losses across groups (p<0.01). Copy number gains and losses were identified in chromosomes 22q, 5q, 7q, 1q, 8q, 20q, 8q, 3p. Note that ER−PR+ differs from other groups in the distribution of copy number gains and losses.

To identify chromosomal regions that may correlate with improved OS by ER/PR categories and hence chemosensitivity, we conducted chromosomal segment analysis. There were 1398 differentially altered genomic segments across all groups. After removing overlapping regions, 191 unique segments remained (**Sup. S12**). The top 20 differentially altered segments were plotted based on the p-value (p<0.05) (**Fig. 5B**). The most significant differentially genomic alteration included 22q11.22 which showed copy loss in 67% (4/6) of ER−/PR+ cases (p=0.000023). Segment 22q11.22 includes the genes IGL, IGLV3-1, MIR5571, IGLL5, IGLJ1. Other differences across four subgroups occurred in segment 5q.21.1 which includes the following genes: SLCO4C1, RN7SKP68, SLCO6A1. We found 33% (2/6) of ER− PR+ cases that presented copy number gains in 5q.21.1 compared to copy number losses found in all other groups (CN losses: 40% (4/10) ER+PR+, 61% (19/31) ER+PR− and 71% (12/17) ER−PR−, p=0.00021). In segment 7q22.3, ER−PR− group presented 29% (5/17) copy losses compared to copy gains in all other groups (CN gains: 10% (1/10) ER+PR+, 39% (12/31) ER+PR− and 50% (3/6) ER−PR+, p=0.0004). In segment 8q24.21 the ER− PR+ group presented 33% (2/6) copy losses compared to copy gains seen in other groups (CN gains: 90% (9/10) ER+PR+, 81% (25/31) ER+PR−, 94% (16/17) ER−PR−, p=0.00119) (**Fig. 5B**). Genes included in other differentially altered segments are reported in **supplementary file 1 (Sup. SF1**).

## Discussion

Previously published data showed the ER−PR+ phenotype predicts favorable tumor biology and long-term survival compared to ER+PR− tumors ^6, 9, 45, 46^. In this study, we show evidence that hormone receptor status impacts the transcriptional programs of HGSC, specifically, univariate analysis of PR+ tumors confer an overall survival benefit with gene set enrichments found in stem cell, immune cell processes and growth and cellular proliferation pathways. These observations suggest hormonal receptor status should be explored further to understand its clinical implications.

The observed decrease in PR+ cells from FTE to STIC to HGSC, along with the majority of our HGSC cohort having negative PR expression, demonstrate a progressive loss of PR expression in HGSC development. A similar pattern of PR expression and loss was seen in double knock out-BRCA1 mice with HGSC^47^. Ongoing research suggests that progesterone signaling may be primarily involved in tumor initiation and not necessarily in tumor progression^47, 48^, especially in high-risk women^49^. Our data is also consistent with literature showing an absence of PR expression in majority of human HGSC tissue^6^.

Our gene expression data showed that ER−PR+ tumors upregulated *SMOC1* compared to ER+PR− tumors and ER−PR−. Secreted modular calcium-binding protein-1 (*SMOC1*) is a gene that is involved in the development of the reproductive tract and mesonephros differentiation and ectopic *SMOC1* expression decreases colony formation, proliferation, and *in vivo* tumor formation as demonstrated by studies in colorectal cancer cells. ^50, 51^. Furthermore, the group of patients with ER−PR+ had tumors that upregulated *SLC4A4*, a gene that is involved in decreasing proliferation and metastasis in addition to suppressing *KRAS* expression in renal cancer cells^52^. In colorectal cancer, downregulation of *SLC4A4* is associated with significantly worse overall survival^53^. However, in prostate cancer, high expression of SLC4A4 in tumor specimens was significantly correlated with disease progression^54^. These data suggest that ER+PR− cases downregulate tumor suppressor-like genes. HGSC is characterized by significant genomic rearrangements and gene specific copy number gains and losses^55^.

We identified a homologous recombination deficiency (HRD)-like phenotype in ER−PR+ tumors which was enriched for DNA damage related genes. Based on copy number alterations analysis, performed on ER/PR subtypes, a strong emphasis is placed on the early genomic alterations of oncogenes and tumor suppressors that occur in ovarian cancer, with *BRCA1* copy number amplifications found in 57% of ER−PR+ subgroup. Using pathway analysis, we identified that *PR* positive tumors upregulated the homologous recombination and DNA repair pathway. Several studies have demonstrated that patients with HR-deficient cancers, especially those with germline *BRCA1/2* mutations, exhibit significantly improved overall survival compared to patients with non-*BRCA* mutated tumors^56^. This effect is more pronounced in *BRCA2* mutation carriers who exhibit even longer survival compared to *BRCA1* carriers^57^. Cyclical use of hormone replacement therapy confers a higher risk of ovarian cancer^58^ compared to the continuous use of estrogen or progestin after menopause in both non-carriers and *BRCA* mutation carriers^59^. These data suggest an improvement in overall survival and *PR* positivity in ovarian cancer tumors could be mediated through homologous recombination pathways.

Gene set enrichment analysis showed that ER−PR+ tumor subtype was associated with the downregulation of immune pathways involved in both innate and adaptive immunity. This feature is consistent with PR’s anti-inflammatory role. Various models, including the incessant ovulation hypothesis propose that chronic exposure to cytokines, chemokines and reactive oxygen species cause DNA damage to the fallopian tube epithelium ^60, 61^. The risk of malignant transformation of distal (secretory) FTE is increased by an altered balance of pro-versus anti-inflammatory signaling molecules during the postovulatory luteal phase ^62^. When progesterone binds to its receptor, it acts as a transcription factor to induce the expression of proteins that dampen inflammation, inhibit cell proliferation, and stimulate repair ^9, 63^. Quantification of a subset of immune cell populations in HGSC across the ER/PR subtypes showed that hormonal status and tumor infiltrating lymphocyte and macrophage presence were not significantly correlated.

Cells exposed to progesterone inhibit inflammatory processes and can decrease the level of macrophage and dendritic cell activation^63–65^. Our results suggest that ER−PR+ tumor cells may confer a better overall survival compared to the other ER/PR subtype patients since an increase in progesterone and its receptors is shown to exert protective effects by decreasing the growth-promoting effects of estrogen, and inducing cell differentiation and apoptosis^66 67^. A corollary response to increased progesterone is to promote survivability of the patient by decreasing the functional activity of the tumor.

While the immune system confers a survival advantage to patients with ovarian cancer, it can also influence the pathogenesis of the disease through the release of cytokines and chemokines^68^. Our transcriptional data of the PR+ tumors showed immune system pathways are downregulated with no significant difference in selected immune infiltration by IHC across ER/PR subtypes. In this study we showed that ER+ tumors have higher levels of macrophages compared to ER− tumors, and ER expression is associated with unfavorable prognosis. This is consistent with data showing that the ER-alpha receptor is expressed in human macrophages and receptor expression can be induced by estradiol (E2)^69^. It is well documented however, that within the tumors, macrophages upregulate ER-alpha expression^70^. In ovarian cancer, macrophages have been shown to display an immunosuppressive and pro-tumoral phenotype, which facilitate chemoresistance and metastasis^71, 72^.

The results of this study may have important implications for the clinical management of patients with HGSC, including the immediate use in patient counseling regarding survival. Our findings emphasize the critical role of PR status relative to ER status based on transcriptional and survival data for high grade serous tumors. The largest retrospective analysis on this subject to date included more than 2000 patients from the Ovarian Tumor Tissue Analysis Consortium. In agreement with our findings, Sieh et al. reported that progesterone receptor status but not estrogen receptor status is associated with improved survival in high grade serous carcinoma^6^. This study further adds to existing literature by exploring transcriptional and genomic phenotypes associated with ER/PR status in human HGSC tumors. A limitation in our study is that our cohort had a relatively low optimal debulking rate (53%) which may suggest bias in the survival analyses. However, the debulking rates were distributed evenly across groups and neoadjuvant chemotherapy use in disease treatment has risen overtime. It will be interesting to see if ER and/or PR expression changes from the chemonaïve to the interval debulked tumor; and if so, to which ER/PR sub-group, and what are the biologic implications of tumor responsiveness to platinum/taxane. Given the important prognostic information provided by hormonal receptor status from this study, patients could be stratified based on their ER/PR status for which precision and targeted therapy will be useful. Multiple phase II trials have shown that ER expression is associated with sensitivity to hormonal therapy^73–75^. However, there are limited available data on treatment response of PR+ tumors to hormonal and other therapies^76^. The distinct phenotypes of high-grade serous carcinoma tumors with observed variable ER/PR expression in the present study warrants further investigation using larger clinical cohorts to identify subpopulations with improved treatment response.

## Data Availability

Clinical data are available upon reasonable request to the authors.
Gene expression array data is available under GSE10971 and GSE28044

https://www.ncbi.nlm.nih.gov/geo/query/acc.cgi?acc=GSE10971

https://www.ncbi.nlm.nih.gov/geo/query/acc.cgi?acc=GSE28044

## Author Contributions

**Conception and design:** L. Dodds, R. Sowamber, A. Milea, P. Shaw, S. George

**Development of methodology:** R. Sowamber, L. Dodds, A. Sanchez-Covarrubias, M. Schlumbrecht, S. George

**Acquisition of data (provided animals, acquired, and managed patients, provided facilities, etc.):** A. Pinto, P. Shaw, S. George

**Analysis and interpretation of data (e.g., statistical analysis, biostatistics, computational analysis):** All authors

**Writing, review, and/or revision of the manuscript:** All authors

**Administrative, technical, or material support (i.e., reporting or organizing data, constructing databases):** L. Dodds, R. Sowamber, A. Sanchez-Covarrubias, S. George

**Study supervision:** S. George

## Acknowledgments

We thank the SCCC Biospecimen Shared Resource, Biostatistics and Bioinformatics Shared Resource and UHealth Department of Pathology for immunohistochemistry, UHN Cancer Biobank Core Laboratory, and the UHN Pathology Research Program. This study was funded by the CDMRP Ovarian Cancer program (W81WH-0701-0371, W81XWH-18-1-0072), Hearing Ovarian Cancer Whispers and the Sylvester Comprehensive Cancer Center. Research reported in this publication was supported by the National Cancer Institute of the National Institutes of Health under Award Number P30CA240139. The content is solely the responsibility of the authors and does not necessarily represent the official views of the National.

