## Supplementary material for "Hormone Receptor Expression and Disease Prognosis in High-Grade Serous Ovarian Cancer": Supplement S2.docx

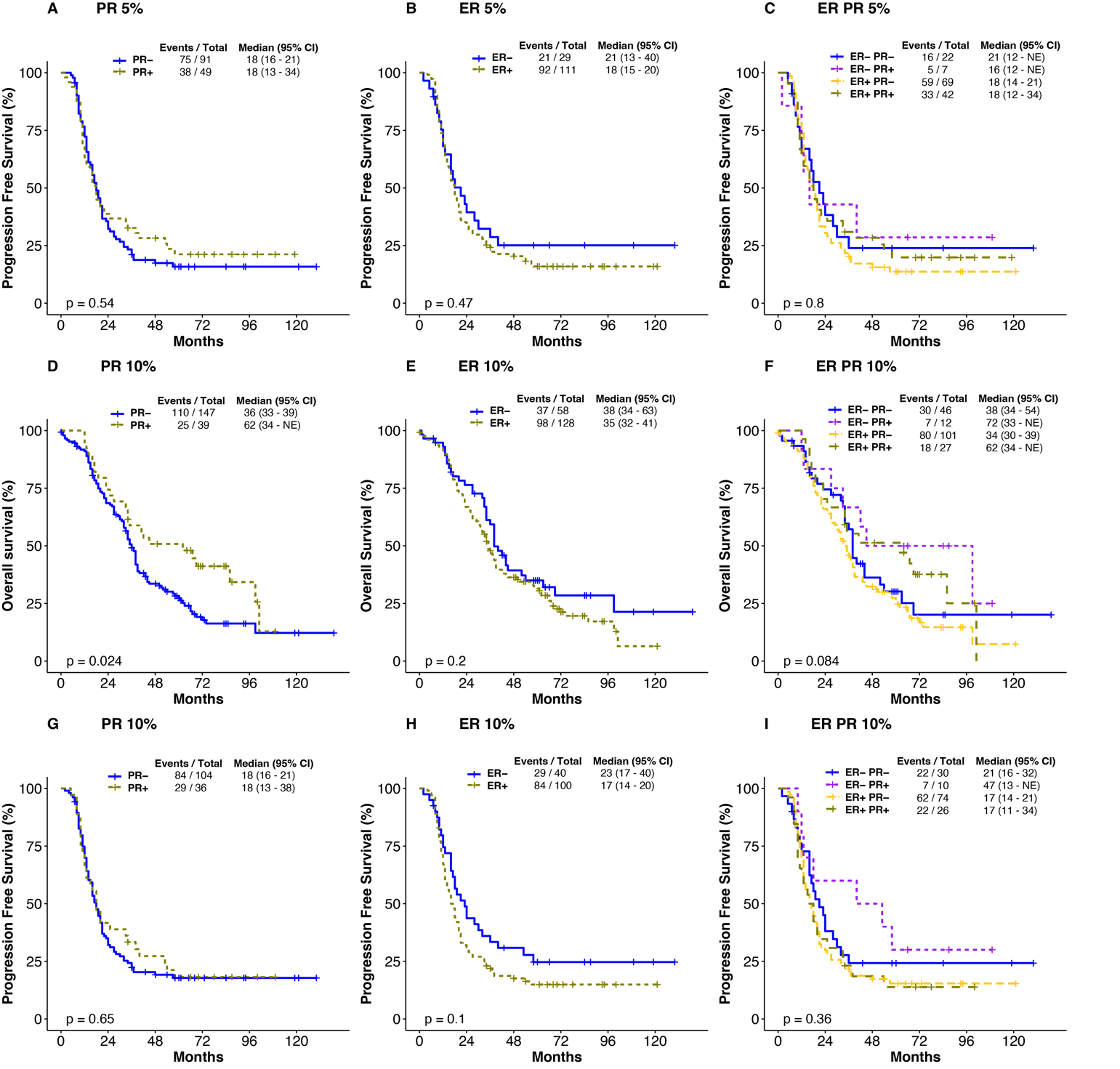

**Supplement S2**: Progression free survival and overall survival in months for ER and PR positive cases at 10% cut-off and 50% cut-off. **A-C**. Progression free survival at ER/PR expression cut-off of 5%. **D-F**. Overall survival for ER/PR subgroups identified at a 10% expression level cut-off. **G-I.** Progression free survival plots for ER/PR subgroups identified at a 10% ER/PR expression cut-off.
