## Supplementary material for "Hormone Receptor Expression and Disease Prognosis in High-Grade Serous Ovarian Cancer": Supplement S3.docx

**Supplementary Table 3**. Cox regression analysis of overall survival at ER/PR 5% expression cutoff.

| **Variable** | **Univariable analysis** | | **Multivariable analysis** | | | |
| --- | --- | --- | --- | --- | --- | --- |
|  |  | | **Model 1**  **(80 events in n=116)** | | **Model 2**  **(103 events in n=141)** | |
|  | **HR (95%CI)** | **p** | **HR (95%CI)** | **p** | **HR (95%CI)** | **p** |
| **Age, 1-year increase** | 1.02 (1.01 – 1.04) | **0.003*** | 1.01 (0.99 – 1.03) | 0.4 | -- |  |
| **Debulking Status** |  |  |  |  |  |  |
| Optimal | Ref |  | Ref |  |  |  |
| Suboptimal | 2.21 (1.54 – 3.17) | **<.0001** | 1.52 (0.92 – 2.50) | 0.10 | 1.87 (1.22 – 2.84) | **0.004*** |
| **Family history of Breast/Ovarian Cancer** |  |  |  |  |  |  |
| Yes | Ref |  | Ref |  | -- |  |
| No | 1.55 (1.03 – 2.33) | **0.036*** | 1.13 (0.71-1.82) | 0.6 | -- |  |
| **Chemotherapy** |  |  |  |  |  |  |
| Platinum + Taxane | Ref |  | Ref |  | Ref |  |
| Platinum only | 1.75 (1.01 – 3.07) | **0.049*** | 3.29 (1.40 – 7.72) | **0.006*** | 2.40 (1.29 – 4.43) | **0.005*** |
| **Number of cycles** |  |  |  |  |  |  |
| ≥6 | Ref |  | Ref |  | Ref |  |
| <6 | 5.24 (2.78 – 9.85) | **<0.001*** | 2.53 (1.02 – 6.30) | **0.046*** | 2.94 (1.38 – 6.29) | **0.005*** |
| **Stage** |  |  |  |  |  |  |
| I – II | Ref |  | Ref |  | -- |  |
| III – IV | 3.14 (1.52 – 6.46) | **0.002*** | 1.30 (0.72 – 2.37) | 0.4 | -- |  |
| **Estrogen Receptor** |  |  |  |  |  |  |
| Positive | Ref |  | -- |  | -- |  |
| Negative | 0.83 (0.54 – 1.27) | 0.4 | -- |  | -- |  |
| **Progesterone Receptor** |  |  |  |  |  |  |
| Positive | Ref |  | Ref |  | Ref |  |
| Negative | 1.69 (1.15 – 2.48) | **0.008*** | 1.30 (0.72 – 2.37) | 0.4 | 1.26 (0.80 – 1.99) | 0.3 |
| **ER/PR combinations** |  |  |  |  |  |  |
| ER+ / PR+ | Ref |  | -- |  | -- |  |
| ER+ / PR- | 1.76 (1.16 – 2.66) | **0.008*** | -- |  | -- |  |
| ER- / PR+ | 0.83 (0.32 – 2.13) | 0.7 | -- |  | -- |  |
| ER- / PR- | 1.33 (0.76 – 2.31) | 0.3 | -- |  | -- |  |

*Statistically significant (p-value <0.05)
