## Supplementary material for "Hormone Receptor Expression and Disease Prognosis in High-Grade Serous Ovarian Cancer": Supplement S4.docx

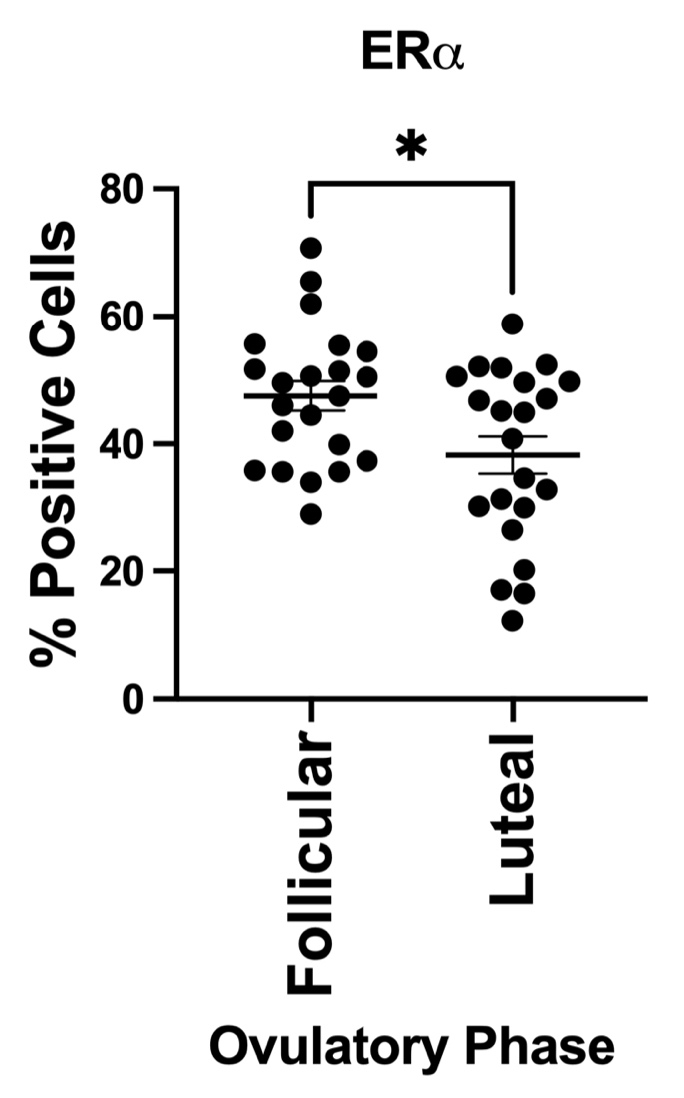

**Supplement S4**: Immunohistochemistry performed on fallopian tube epithelial tissues of the luteal (n=22) and follicular phases (n=22) shows significantly higher expression of estrogen receptor in tissue with follicular phase status (p=0.01).
