## Supplementary material for "Hormone Receptor Expression and Disease Prognosis in High-Grade Serous Ovarian Cancer": Supplement S6.docx

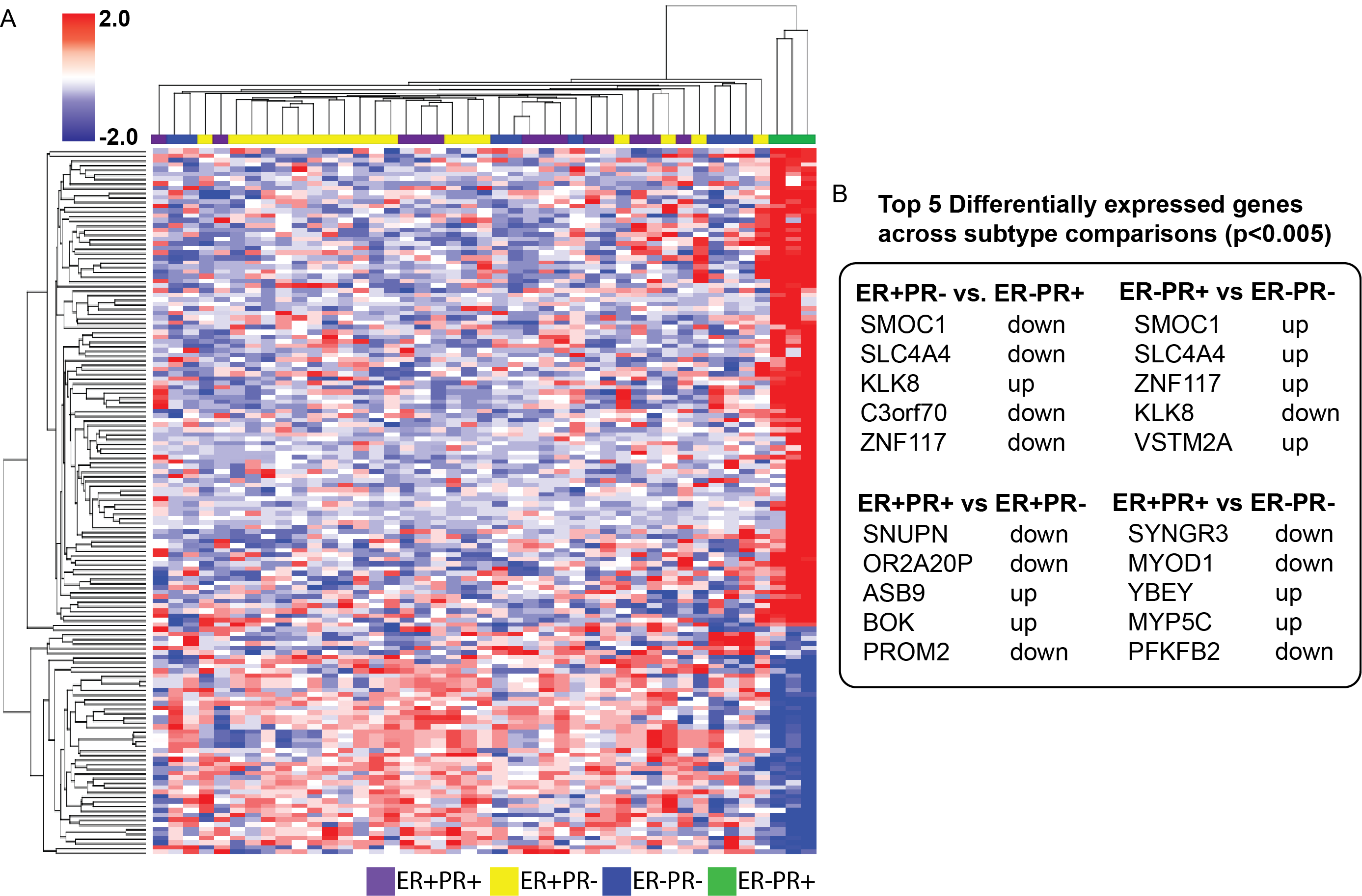

**Supplement S6**: Differentially expressed genes between different combinations of ER/PR expression groups. A. Heatmap showing unsupervised hierarchical clustering of differentially expressed genes between subgroups, there is a clear clustering of ER-PR+ tumors. B. Top 5 differentially expressed genes identified across selected ER/PR comparisons (p<0.05); the genes SMOC1, SLC4A4, KLK8 and ZNF117 where present whenever ER-PR+ was selected as one of the comparison groups.
