## Supplementary material for "Hormone Receptor Expression and Disease Prognosis in High-Grade Serous Ovarian Cancer": Supplement S7.docx

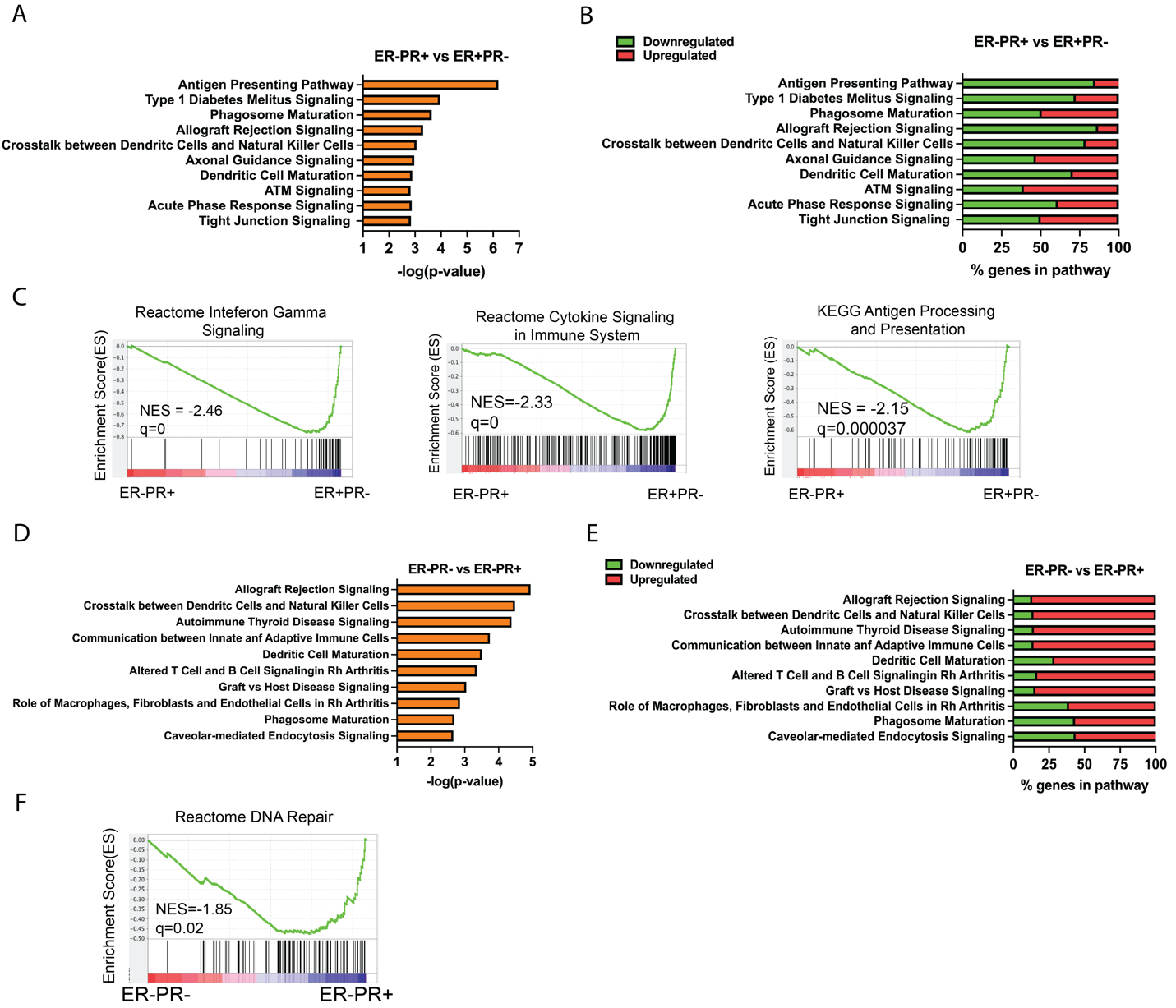

**Supplement S7:** GSEA pathway analysis across selected ER/PR groups. **A.** Comparison between ER-PR+ vs ER+PR- show negative enrichment of immune gene sets**. B.** Percentage of genes in a pathway for the same group comparison in A. **C.** Visual representation of GSEA enrichment plots for a comparison between ER-PR+ and ER+PR- cases shows enrichment for interferon gamma signaling (NES= -2.46, q=0), cytokine signaling in the immune system (NES = -2.33, q=0) and antigen processing and presentation (NES = -2.15, q=0.000037). **D.** Comparison between ER-PR- vs ER-PR+ groups show pathways regulated solely by PR. **E.** Percentage of genes in a pathway for the same group comparison in D. **F.** In the absence of ER, PR+ tumors upregulate the reactome DNA repair pathway.
