## Supplementary material for "Hormone Receptor Expression and Disease Prognosis in High-Grade Serous Ovarian Cancer": Supplement S11.docx

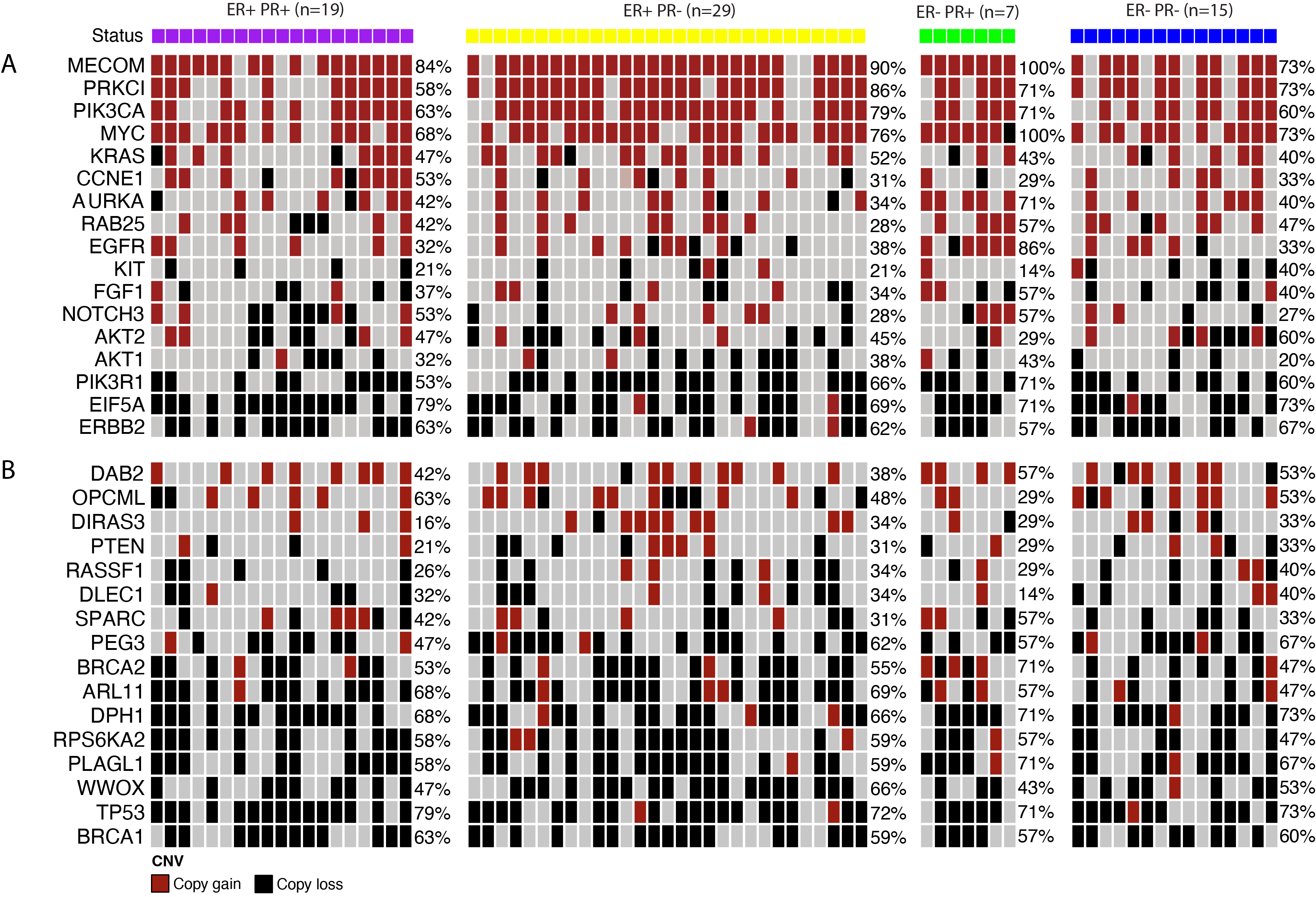

**Supplement S11**. Copy number variation gene analysis in a subset of genes known in ovarian cancer pathogenesis in TCGA. **A**. Oncogenes associated with epithelial ovarian cancer **B.** Putative tumor-suppressor genes in epithelial ovarian cancer.
